# Autophagy inhibitors block pathogenic NET release in immune-mediated inflammatory disease without impairing host defence

**DOI:** 10.1101/2025.06.23.25330105

**Authors:** Andy Nolan, Daniel M Foulkes, Emma E Fairweather, Michele Fresneda Alarcon, Christina Linford, Andrew Sellin, Zoe McLaren, Patrick A Eyers, Helen Wright

## Abstract

**Objectives:** Activation of neutrophils and release of neutrophil extracellular traps (NETs), proteases and reactive oxygen species (ROS) is pathogenic in immune-mediated inflammatory diseases like rheumatoid arthritis (RA) and systemic lupus erythematosus (SLE), driving inflammation and damaging host tissues. The aim of this research was to identify small molecule inhibitors of NET production using a highly-curated panel of narrow-spectrum small molecule kinase inhibitors termed the Kinase Chemogenomic Set (KCGS).

**Methods:** Neutrophils were isolated from healthy controls (HC) and people with RA or SLE. Over 220 small molecules were screened for their ability to inhibit NET production and decrease ROS production without impairing neutrophil apoptosis or killing of *S. aureus* bacteria.

**Results:** Fifty compounds, nominally targeting 29 individual protein kinase targets, were found to inhibit NOX2-dependent (PMA-stimulated) and NOX2-independent (A23187-stimulated) NET production (p<0.05, n=5 HC, RA and SLE). Of these, seven compounds did not significantly impair ROS production or apoptosis. The deconvoluted targets of these small molecules inhibit kinases that operate in three cellular pathways: autophagy, cell cycle checkpoint and epidermal growth factor (EGF) tyrosine kinase signalling. Of these, only inhibitors of ULK1, JNK and ROCK1/2, broadly implicated in the regulation of autophagy, did not significantly impair bacterial killing (n=5 HC, p>0.05). Autophagy inhibitors were also able to inhibit immune-complex driven NET production (p<0.05, n=5 HC, RA and SLE).

**Conclusion:** We propose that autophagy signalling pathways represent novel and exciting targets for the development of small molecule therapeutics to block unwanted neutrophil activation and NET release in immune-mediated inflammatory disease.

## Introduction

Neutrophil extracellular traps (NETs) are DNA:protein complexes that are expelled from neutrophils in response to infectious or inflammatory stimuli. Significant evidence supports the role of externalised, double-stranded DNA and post-translationally modified proteins on NETs in driving immune-mediated inflammation [1-3]. Biologically-active serine proteases externalised on NETs, including neutrophil elastase and collagenase (MMP8), cause damage to local tissues during episodes of immune-mediated inflammation [3, 4]. NETs are present in RA blood smears, in RA synovial tissues and in kidneys of people with SLE, driving localised inflammation and the development of auto-immunity [5-7]. Many people with RA and SLE do not achieve good disease control with front-line disease modifying anti-rheumatic drugs (DMARDs). The alternative is treatment with biologics such TNF inhibitors or JAK inhibitors for RA, and belimumab (anti-BAFF/BLyS) for SLE. Rituximab, which targets CD20^+^ B-cells, may be prescribed for both RA and SLE patients, however none of these therapies directly target inflammation driven by neutrophils. Neutrophils are one of the few immune cells with the cytotoxic potential to inflict damage to host tissues, as well as drive inflammation through NET release. Our screening approach converges on several kinase targets whose inhibition interferes with NET production, and we therefore believe neutrophils are a novel and exciting target for further kinase inhibitor investigation and drug development.

## Methods

### Ethics

This study was approved by the University of Liverpool Central University Research Ethics Committee C (Ref: 10956), and NRES Committee North West (Greater Manchester West, UK) (Ref: 11/NW/0206). All participants gave written, informed consent in accordance with the declaration of Helsinki. All patients with clinician-diagnosed RA or SLE were recruited from Liverpool University Hospital NHS Foundation Trust. Healthy controls were recruited from staff at the University of Liverpool. All participants were over the age of 18 years and free of infection.

### Neutrophil isolation

Ultrapure human neutrophils (>99%) were isolated from heparinised peripheral blood within 1h of venipuncture using Hetasep and the Easysep human neutrophil isolation kit (Stem Cell). Neutrophils were resuspended in RPMI 1640 media (Life Technologies) containing L-glutamine (2mM) and Hepes (25mM).

### Cell Signalling Modulators

The KCGS library of 187 narrow-specificity protein kinase/pseudokinase inhibitors [8, 9] targeting >200 protein kinases was purchased from cancertools.org. Compounds (5 μM) were initially screened in high-throughput neutrophil assay format. Thirty-one separate FDA-approved kinase inhibitors (Selleckchem) were also included at a final concentration of 10 μM (Supplementary Table 1). Staurosporine, a well-known multi-kinase inhibitor which induces apoptosis, was included as a positive control (1 μM). Other commercially-available signalling pathway inhibitors were purchased from Abcam, MedchemExpress, Selleckchem and Tocris, and used at 5 μM. Small molecules (typically 500 nL) were dispensed into 96 well plates using an Echo 650 acoustic liquid handler (Beckman Coulter) prior to neutrophil addition.

### Measurement of NET Production with Sytox Green

Neutrophils (10^5^ in 100μL) were seeded in black 96-well plates containing Sytox green reagent (5mM) in the presence or absence of compounds (0.5% DMSO v/v) for 15 min at 37^°^ C. Subsequently, neutrophils were stimulated with phorbol 12-myristate 13-acetate (PMA, 600 nM), A23187 (5 mM) or antibody immune complexes (10 % w/v) [10] and incubated for up to 4h at 37 ^o^ C, prior to fluorescence measurements using a Fluostar Optima plate reader (excitation 488nm, emission 530nm).

### Imaging of neutrophils on cover slips

Neutrophils (2×10^5^ cells/500 μL) were seeded in RPMI medium in a 24-well plate containing poly-L-lysine coated coverslips [1]. Cells were pre-treated with compounds for 15 min and allowed to adhere for 30 min prior to stimulation with PMA (600 nM). Cells were incubated for a further 4h to permit NET production. Cells adhered to coverslips were fixed with 4% paraformaldehyde prior to staining. Primary antibodies were mouse anti-myeloperoxidase (1:1000, Abcam) and rabbit anti-elastase (1:200, Abcam). Secondary antibodies used were anti-rabbit AlexaFluor488 and anti-mouse AlexaFluor647 (1:2000, Life Technologies). Coverslips were washed prior to staining with DAPI (1μg/mL). Slides were imaged on an LSM800 microscope (Zeiss) using a 20X objective.

### Detection of ROS Production by Luminol-Enhanced Chemiluminescence

Neutrophils (10^5^) were resuspended in 200 mL HBSS containing luminol (10 mM). ROS production was stimulated with PMA (600 nM) and monitored continuously for 1h in a Fluostar Optima plate reader at 37 °C.

### Measurement of Apoptosis by flow cytometry

Following incubation +/-compounds for 24h, 5×10^4^ neutrophils were removed from culture and diluted with 200 mL HBSS containing 0.5 mL Annexin V-488, and incubated at room temperature in the dark for 15 min. HBSS containing propidium-iodide (final concentration of 1 mg/mL) was added to total volume of 250 mL and samples were immediately analysed using an EasyCyte Guava flow cytometer (5,000 events per sample).

### Bacterial killing assay

Bacterial killing assay was performed as previously described [11]. Briefly, live *S. aureus* were washed, suspended at 5×10^8^/mL in HBSS and opsonised with 10% human AB serum (Merck) for 30 min at 37°C. Freshly-isolated neutrophils (10^6^/mL) were primed with TNF*α* (10 ng/mL) and incubated for 90 min at 37°C with gentle agitation and serum-opsonised bacteria at a ratio of 1:10. Neutrophils were lysed to release live bacteria by serial dilution in distilled water and vigorous vortexing, before plating on LB agar and overnight incubation at 37 °C. Colonies were counted and results calculated as percentage of bacteria killed compared to bacteria only (no neutrophils) samples.

### Statistical analysis

Univariate analysis of experimental data was carried out using ANOVA in R (v4.0.2).

## Results

### NET production is regulated by kinase signalling

To evaluate the role of kinase-based signalling in the regulation of NET production (NETosis) by human neutrophils we first incubated healthy control neutrophils (HC, n=4) with the non-specific kinase inhibitor staurosporine (1 μM) for 15 min prior to stimulation of NETosis by PMA (NOX2-dependent) or A23187 (NOX2-independent). Staurosporine significantly inhibited NETosis at 4h (Figure 1A, p<0.01). We next performed screening of ∼220 compounds including the KCGS library and compounds from an in-house FDA-approved drug library. NET inhibition ranged from no inhibition to almost complete inhibition, both after PMA-induced and A23187-induced NET induction (Figure 1B). Fifty compounds in the screen were found to significantly inhibit PMA- and A23187-NETosis at 4h (p<0.05, n=4).

**Figure 1.**
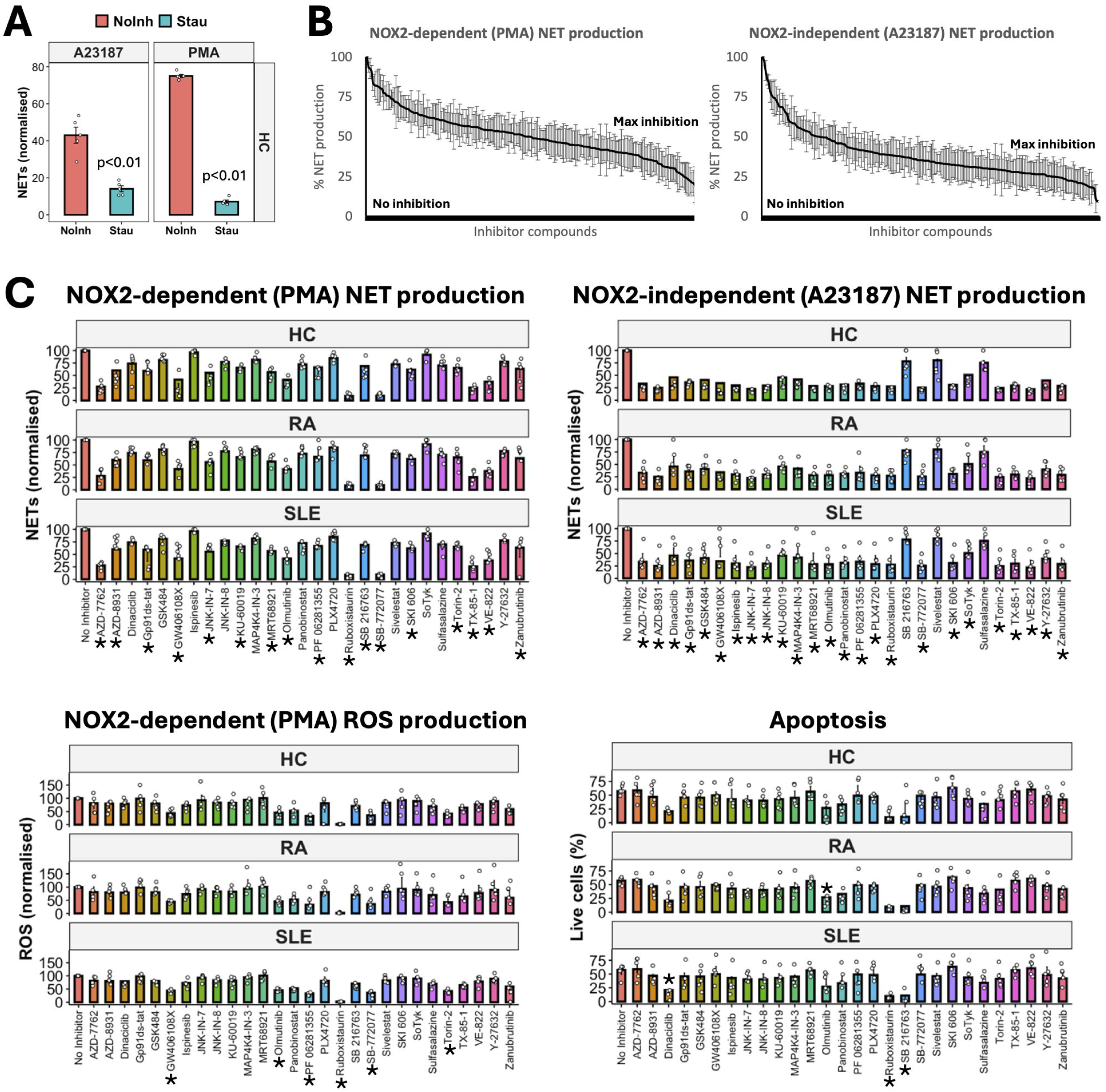
Kinase inhibitors block neutrophil extracellular trap (NET) production. (A) NET production in response to A23187 (NOX2-independent) and PMA (NOX2-dependent) is blocked by Staurosporine (Stau, p<0.01, n=4). (B) Library screen (KCGS, FDA) to identify compounds that significantly inhibit NET production (n=4). (C) Heathy (HC), rheumatoid arthritis (RA) and systemic lupus erythematosus (SLE) neutrophil functional responses after compound exposure. NET production, ROS production and apoptosis (20h) were measured. (* p<0.05 in 2/3 groups, each group n=5).

The most-likely molecular kinase target(s) of each of the narrow-spectrum 50 KCGS/FDA compounds was assessed according to published data [8] and data deconvoluted to generate a condensed list of 29 compounds with shared high-likelihood targets (Supplementary Table 2). The effects of each compound on neutrophil ROS production (which is critical for host defence) was measured using HC, RA and SLE neutrophils (n=5 each group) alongside assessment of cell viability (Figure 1C). Where relevant, structurally diverse compounds targeting the same kinases, identified from the KCGS/FDA libraries, were purchased to validate and extend our datasets. Seventeen compounds (∼60%) significantly inhibited PMA-induced NETosis (p<0.05), and 25 compounds (86%) significantly decreased A23187-induced NETosis in at least two of the three participant groups (p<0.05, Figure 1C). Notably, ROS production was inhibited by the compounds GW406108X (Kif5/Kin12 inhibitor), Olmutinib (EGFR), PF 06281355 (a myeloperoxidase inhibitor), Ruboxistaurin (LY333531, an orally-active PKC-β inhibitor), SB-772077 (ROCK inhibitor) and Torin-2 (mTOR/DNA-PK inhibitor) in at least two of three participant groups (p<0.05). In addition, Ruboxistaurin significantly enhanced neutrophil apoptosis in all participant groups tested (p<0.01). In contrast, the GSK3 α/β inhibitor SB 216763 significantly enhanced apoptosis in RA and SLE neutrophils (p<0.01), Dinaciclib (a pan CDK inhibitor) enhanced SLE neutrophil apoptosis (p<0.05) whereas Olmutinib (an EGFR tyrosine kinase inhibitor) enhanced RA neutrophil apoptosis (p<0.05). Compounds that significantly enhanced neutrophil apoptosis were not investigated further, since they were considered likely to impair host defence through inhibition of core neutrophil killing properties.

### Autophagy inhibitors prevent NET release but preserve bacterial killing capacity

The kinase targets of the inhibitors that significantly inhibited NETs without significantly impairing ROS or apoptosis fell into groups that regulate core signalling networks related to autophagy, cell cycle checkpoints and EGF tyrosine kinase signalling (Figure 2A). Seven compounds were investigated further to evaluate unwanted effects on host defence mechanisms. All compounds were able to significantly inhibit immune-complex induced NET production in HC, RA and SLE neutrophils (Figure 2B, p<0.05, n=5 each group) with the exception of the pan-EGFR inhibitor AZD-8931, which had no statistical effect in SLE neutrophils (p=0.052). However, four compounds significantly inhibited neutrophil killing of *S. aureus* bacteria, significantly impairing a critical function of neutrophils in the host defence response (p<0.05, Figure 2C, n=5 each group). The compounds GW406108X, which targets Kinesin-12 and ULK1 and is a potent autophagy inhibitor, JNK-IN-7 which targets c-Jun N-terminal kinases (JNK), and SB-772077 which is a ROCK1/2 inhibitor, did not significantly impair bacterial killing. The ability of these three compounds to inhibit NET production was independently confirmed by immuno-fluorescence staining of NOX2-induced NETs on coverslips (Figure 2D).

**Figure 2.**
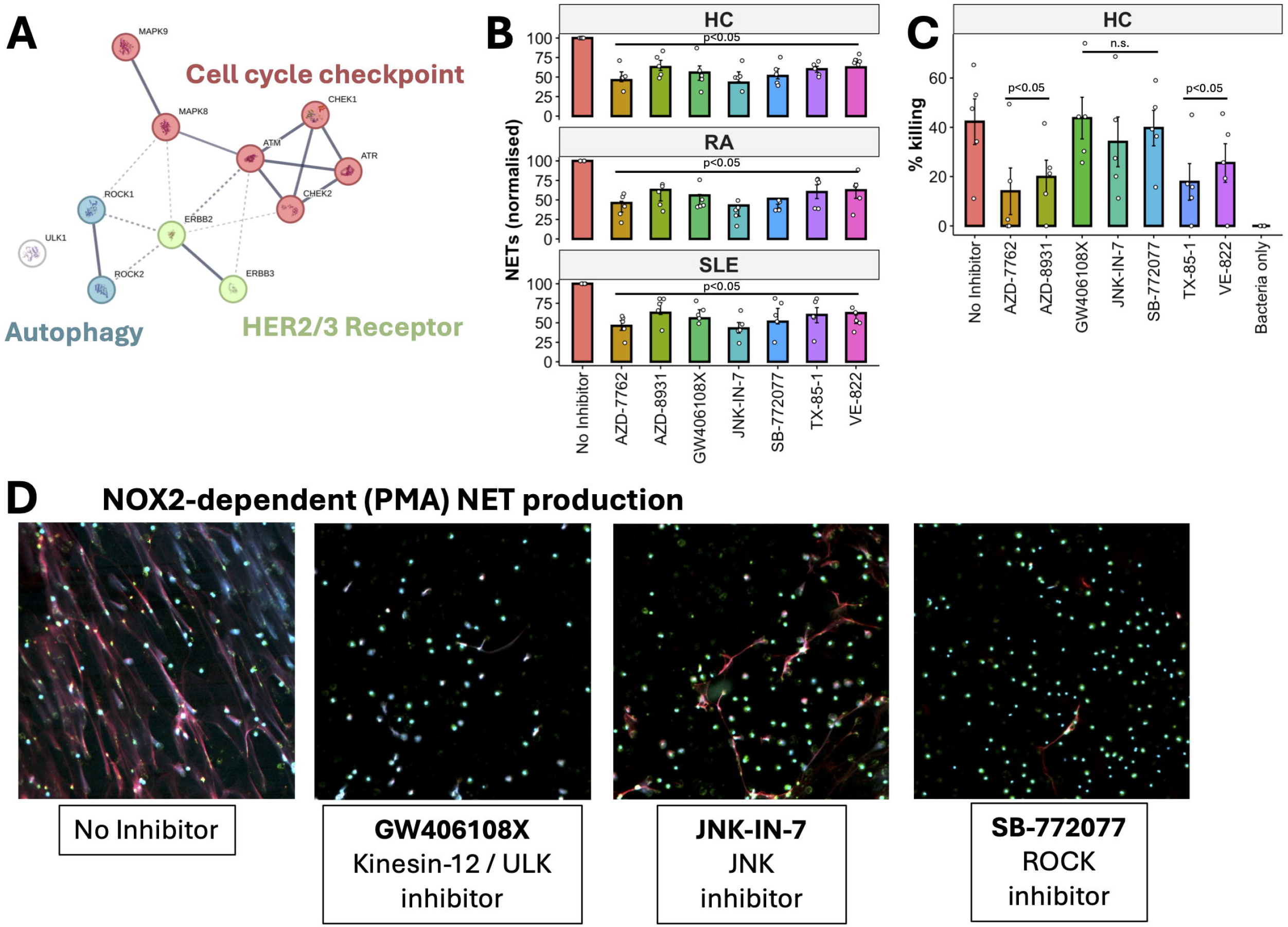
Kinase inhibitors block NET production without impairing host defence. (A) STRING network of compound targets that regulate NET production without impairing ROS production or apoptosis. (B) Small molecules significantly decrease NET production in response to immune-complexes (p<0.05). (C) Effect of kinase inhibitors on neutrophil killing of S. aureus (p<0.05, n.s. = not significant). (D) Inhibition of PMA-stimulated NET production by compounds GW406108X, JNK-IN-7 and SB-772077 after 4h. NETs are stained for DNA (DAPI, blue), MPO (red), neutrophil elastase (green).

## Discussion

In this study we carried out screening experiments to identify subsets of target-validated kinase inhibitors that can inhibit pathogenic NET production in RA and SLE, without significantly impairing neutrophil host defence functions or neutrophil life span. Several signalling pathways regulating NETosis have been proposed, including Raf-MEK-ERK [12], RIPK1-RIPK3-MLKL [13, 14] and ERK, AKT, p38-MAPK and cSrc [15]. Cell cycle firing at the G1/S transition and the activation of CDK4 and CDK6 has also been identified as essential to the initiation of NETosis [16]. Our initial experiments suggested key roles for several cell cycle checkpoint proteins in regulating NETosis, however exposure to inhibitors of CHEK1/2 and ATM/ATR led to significant impairment of neutrophil bacterial killing capacity, which potentially makes them unsuited as NET inhibitors. We also identified epidermal growth factor tyrosine kinase signalling via the HER2/3 receptor (ERBB2/ERBB3) as a regulator of NETosis. This receptor complex is targeted in solid tumour cancers, especially HER2-positive breast cancer, by the drug Herceptin. However, in our experiments we found that inhibition of these tyrosine kinase receptor classes also significantly impaired bacterial killing.

Our screen of narrow-spectrum kinase inhibitors and associated compounds led us to identify a short-list of three candidate small molecule inhibitors that appear to target kinases that are known to operate in autophagy-based signalling pathways as modulators of NETosis. Compound GW406108X targets ULK1 which is a critical regulator of autophagy initiation and autophagosome formation [17]. Both JNK (inhibited by JNK-IN-7) and ROCK1/2 (inhibited by SB-772022) initiate autophagy in response to oxidative and metabolic stress *via* Beclin-1, a master regulator of autophagosome assembly [18-20]. Autophagy is a key pathway regulating organelle recycling and cell death, and is a known regulator of neutrophil activation associated with damage to host tissue during aberrant activation, including degranulation, ROS production and NETosis [21]. Activation of autophagy in neutrophils is independent of NOX2 activation, with neutrophils from patients with chronic granulomatous disease (lacking NOX2) still being able to undergo autophagy in response to PMA [22]. Inhibition of autophagy prevents the decondensation of intracellular chromatin, an important step in NET production [22]. Interestingly, autophagy inhibition was also shown to block NETosis in response to oligomeric silsesquioxane nanoparticles, structures which are endocytosed by neutrophils in a similar way to auto-immune complexes [23]. However, it should be noted that these studies employed wortmannin to inhibit autophagy, a non-specific phosphatidylinositol 3-kinases (PI3Ks) and phosphatidylinositol 3-kinase-related kinases (PIKKs) inhibitor, which is likely to inhibit multiple cellular processes in addition to autophagy.

In conclusion, pathogenic NETosis is an attractive and novel candidate for targeting new therapeutics in auto-immune disease, since NET release is directly responsible for both the initiation of auto-immunity [1, 2, 24] and for directing immune-mediated damage to host tissues, including synovial tissues and cartilage in RA [2, 6, 25, 26] and blood vessels in SLE [7, 27-29]. Inhibition of NETs in the collagen-induced arthritis RA model reduces disease severity by lowering cytokine secretion by macrophages [30]. In an SLE-mouse model, NET inhibition decreases skin, kidney and vascular damage [31]. Therefore, modulation of NETosis is considered an important pharmacological goal in a variety of immune-system disorders. Using an unbiased high throughout screen, we have identified three small molecule inhibitors of autophagy-related pathways that block pathogenic NET production by RA and SLE neutrophils without significantly impairing host defence. We propose that autophagy represents a novel and exciting new signalling target for the development or repurposing of therapeutics [32] that block unwanted neutrophil activation in immune-mediated inflammatory disease.

## Acknowledgements

We thank the clinical staff at Liverpool University Hospitals NHS Foundation Trust for help in recruiting participants to the study.

## Group authorship list

Andy Nolan performed the experiments, formal analysis, methodology, visualisation, writing – review and editing

Daniel Foulkes methodology, writing – review and editing

Emma Fairweather methodology, writing – review and editing

Michele Fresneda Alarcon performed the experiments, writing – review and editing

Christina Linford performed the experiments, formal analysis, methodology, writing – review and editing

Andrew Sellin methodology, writing – review and editing

Zoe McLaren conceptualisation, funding acquisition, participant screening and recruitment,

writing – review and editing

Patrick Eyers conceptualisation, funding acquisition, methodology, supervision, writing –

review and editing, writing – review and editing

Helen Wright conceptualisation, data curation, formal analysis, funding acquisition, methodology, project administration, supervision, visualisation, writing – original draft, writing – review and editing

## Funding

This research was funded by a Connect Immune Research and The Lorna & Yuti Chernajovsky Biomedical Research Foundation Grant (Grant No. 22925).

## Conflict of interest statement

None

## Data availability statement

All data are available from the corresponding author upon reasonable request.

## Notes

### Competing Interest Statement

The authors have declared no competing interest.

### Author Declarations

This study was approved by the University of Liverpool Central University Research Ethics Committee C (Ref: 10956), and NRES Committee North West (Greater Manchester West, UK) (Ref: 11/NW/0206). All participants gave written, informed consent in accordance with the declaration of Helsinki.

