## Supplementary Table for "Autophagy inhibitors block pathogenic NET release in immune-mediated inflammatory disease without impairing host defence"

**^1^Institute of Life Course and Medical Sciences, University of Liverpool, Liverpool UK**

**^2^Department of Biochemistry, Cell and Systems Biology, Institute of Systems, Molecular and Integrative Biology, University of Liverpool UK**

^3^**Liverpool University Hospitals NHS Foundation Trust, Liverpool, UK**

**Supplementary Table 1. Compounds from the FDA approved library that were used to screen for inhibition of NET production.** * gp91-ds-TAT and GSK484 were not part of the FDA library and were purchased from MedchemExpress.

| **Compound Name** | **Characterized Molecular target(s)** |
| --- | --- |
| Acalabrutinib | Bruton’s tyrosine kinase (BTK) |
| Acetylcysteine | Reactive Oxygen Species (ROS) |
| Almitrine mesylate | Calcium-dependent potassium channel |
| Belinostat | Histone deacetylase (HDAC) |
| Casticin | Transforming growth factor beta 1 (TGFβ1) |
| Chloroquine | Autophagy |
| Curcumin | Nuclear Factor kappa-light-chain-enhancer of activated B cells (NF-κB), Histone deacetylase (HDAC), Histone Acetyltransferase, Nuclear factor erythroid 2-related factor 2 (Nrf2) |
| Daphnetin | Protein Kinase A (PKA), Epidermal growth factor receptor (EGFR), Protein Kinase C (PKC) |
| Deucravitinib | Tyrosine kinase 2 (Tyk2) |
| Doxycycline Hyclate | Matrix Metalloproteinase (MMP) |
| Enzastaurin | Protein Kinase C (PKC) |
| Fargesin | Anti-inflammatory |
| Fasudil | Autophagy, Rho-associated protein kinase (ROCK) |
| Glycyrrhizin | Dehydrogenase, Monoamine oxidase (MAO), High-Mobility Group |
| gp91-ds-TAT * | NADPH oxidase assembly peptide inhibitor |
| GSK484 * | Protein arginine deiminase (PAD4) |
| Hydroxychloroquine Sulfate | Autophagy |
| Hypericin | Monoamine oxidase (MAO) |
| Ibrutinib | Bruton’s tyrosine kinase (BTK) |
| Marimastat | Matrix Metalloproteinase (MMP) |
| Methotrexate | Dihydrofolate reductase (DHFR) |
| Olmutinib | Epidermal growth factor receptor (EGFR), Bruton’s tyrosine kinase (BTK) |
| Panobinostat | Histone deacetylase (HDAC) |
| PF 06281355 | Myeloperoxidase (MPO) |
| Quercetin | Anti-inflammatory |
| Ruboxistaurin | Protein Kinase C (PKC) |
| Silvelestat Sodium Salt | Neutrophil elastase |
| Sulfasalazine | Sepiapterin reductase (SPR) |
| Tofacitinib | Janus Kinase 1 (JAK1), Janus Kinase 3 (JAK3) |
| Vorinostat | Histone deacetylase (HDAC) |
| Zanubrutinib | Bruton’s tyrosine kinase (BTK) |

**Supplementary Table 2. Condensed list of commercial compounds that were used to screen for inhibition of NETosis, ROS production and apoptosis.**

| **Compound** | **Molecular Target** |
| --- | --- |
| AZD-7762 | Checkpoint kinase (Chk) |
| AZD-8931 | Epidermal growth factor receptor (Erbb2/Erbb3) |
| Deucravacitinib | Tyrosine kinase 2 (Tyk2) |
| Dinaciclib | Cyclin-dependent kinases (CDK) |
| gp91-ds-TAT | NADPH oxidase (NOX2) |
| GSK484 | Protein arginine deiminase (PAD4) |
| GW406108X | Kinesin-12/ULK1 |
| Ispinesib | Kinesin spindle protein |
| JNK-IN-7 | c-Jun N-terminal kinase (JNK) |
| JNK-IN-8 | c-Jun N-terminal kinase (JNK) |
| KU-60019 | Ataxia-telangiectasia mutated kinase (ATM) |
| MAP4K4-IN-3 | Mitogen-activated protein kinase kinase kinase kinase 4 (MAP4K4) |
| MRT68921 | Unc-51-like autophagy-activating kinases 1 (ULK1) |
| Olmutinib | Epidermal growth factor receptor tyrosine kinase (EGFR) |
| Panobinostat | Histone deacetylase (HDAC) |
| PF06281355 | Myeloperoxidase |
| PLX4720 | B-RafV 600E |
| Ruboxistaurin | Protein kinase C β |
| SB 216763 | Glycogen synthase kinase 3 (GSK3) |
| SB-772077 | Rho-associated protein kinase (ROCK) |
| Sivelestat | Neutrophil elastase |
| SKI606 | Src/Abl |
| Sulfasalazine | NF kappaB |
| Torin-2 | Mammalian target of rapamycin (mTOR) |
| TX-85-1 | Epidermal growth factor receptor (Erbb3) |
| VE-822 | Ataxia telangiectasia and Rad3-related protein (ATR) |
| Y-27632 | ROCK1/ROCK2 |
| Zanubrutinib | Bruton’s tyrosine kinase (BTK) |
